# Characteristics and outcomes of patients with COVID-19 at high-risk of disease progression receiving sotrovimab, oral antivirals or no treatment in England

**DOI:** 10.1101/2022.11.28.22282808

**Authors:** Vishal Patel, Marcus J. Yarwood, Bethany Levick, Daniel C. Gibbons, Myriam Drysdale, William Kerr, Jonathan D. Watkins, Sophie Young, Benjamin F. Pierce, Emily J. Lloyd, Helen J. Birch, Tahereh Kamalati, Stephen J. Brett

**Affiliations:** GSK, Middlesex, UK; Imperial College Health Partners, London, UK; OPEN Health Evidence & Access, York, UK; Department of Surgery and Cancer, Imperial College, London, UK

**Keywords:** COVID-19, molnupiravir, monoclonal antibody, nirmatrelvir/ritonavir, Omicron BA.1, Omicron BA.2, oral antivirals, sotrovimab

## Abstract

**Introduction:** There is limited real-world evidence surrounding the effectiveness of early, mild-to-moderate COVID-19 treatments following the emergence and dominance of Omicron SARS-CoV-2 subvariants. Here, characteristics and acute clinical outcomes are described for patients with COVID-19 treated with sotrovimab, nirmatrelvir/ritonavir or molnupiravir, or patients at highest risk per NHS criteria but who were untreated.

**Methods:** Retrospective cohort study of non-hospitalised patients who received early treatment for, or were diagnosed with, COVID-19 between 1 December 2021 and 31 May 2022, using data from the Discover dataset in north-west London. Patients were included if aged ≥12 years and treated with sotrovimab, nirmatrelvir/ritonavir or molnupiravir, or were untreated but expected to be eligible for early treatment per NHS highest-risk criteria at time of diagnosis. Outcomes were reported for 28 days from COVID-19 diagnosis (index). Subgroup analyses were conducted in patients with advanced renal disease, those aged 18–64 and ≥65 years and by period of Omicron BA.1, BA.2 and BA.5 *(post-hoc* exploratory analysis) predominance.

**Results:** A total of 696 patients prescribed sotrovimab, 337 prescribed nirmatrelvir/ritonavir, 470 prescribed molnupiravir and 4,044 eligible high-risk untreated patients were included. A high proportion of patients on sotrovimab had advanced renal disease (29.3%), ≥3 high-risk comorbidities (47.6%) and were aged ≥65 years (36.9%). In total, 5/696 (0.7%) patients on sotrovimab, <5/337 (0.3–1.2%) patients on nirmatrelvir/ritonavir, 10/470 (2.1%) patients on molnupiravir and 114/4,044 (2.8%) untreated patients were hospitalised with COVID-19 as the primary diagnosis. Similar results were observed across all subgroups and during Omicron subvariant periods.

**Conclusion:** Patients who received sotrovimab appeared to show evidence of multiple comorbidities that may increase risk of severe COVID-19. Low hospitalisation rates were observed for all treated cohorts across subgroups and periods of predominant variants of concern. These descriptive results require confirmation with comparative effectiveness analyses adjusting for differences in underlying patient characteristics.

**Key points:** *Why carry out this study?:*

- There is limited real-world evidence surrounding early, mild-to-moderate COVID-19 treatments, particularly during Omicron subvariant dominance periods, and the UK National Institute for Health and Care Excellence has recommended more is gathered.
- We described patient characteristics and clinical outcomes among patients treated with sotrovimab, nirmatrelvir/ritonavir, molnupiravir or who met the highest-risk eligibility criteria but were untreated.

*What was learned from the study?:*

- Sotrovimab was often utilised amongst more elderly and at-risk patients, such as those with advanced renal disease, than patients treated with nirmatrelvir/ritonavir or molnupiravir.
- We found that hospitalisation rates were low across all treated cohorts.
- For patients treated with sotrovimab, clinical outcomes appeared consistent when observed across the age subgroups and Omicron subvariant periods, as well as among patients with advanced renal disease.

## Introduction

Coronavirus disease 2019 (COVID-19) is caused by infection with severe acute respiratory syndrome coronavirus 2 (SARS-CoV-2), and was initially identified in December 2019.^1^ The rapid global spread of SARS-CoV-2 resulted in the declaration of a pandemic by the World Health Organization in March 2020.^2^ Some patients, such as older individuals or those with advanced renal disease, diabetes, cancer, chronic obstructive pulmonary disease or cardiovascular disease, are at higher risk of developing severe COVID-19, which may result in hospitalisation or death.^3,4^ Moreover, data have suggested that immunocompromised individuals or transplant recipients, who are also considered at high risk of severe COVID-19, have blunted serological responses to vaccines.^5,6^

In England, early, mild-to-moderate COVID-19 treatment with either antivirals or monoclonal antibodies (mAbs) is recommended for people with ‘highest-risk’ conditions, as defined by national clinical guidance. During the study period these included cancer, advanced renal and liver disease, human immunodeficiency virus/acquired immune deficiency syndrome (HIV/AIDS), solid organ and stem cell transplant recipients and rare neurological conditions.^7,8^ Of particular note, it has been recently reported that people with advanced renal disease are at particularly high risk of severe COVID-19 outcomes.^4^

Sotrovimab is a recombinant human IgG1κ monoclonal antibody derived from the parental mAb S309, a potent neutralising mAb directed against the spike protein of SARS-CoV-2,^9-12^ that was shown in a randomised clinical trial to significantly reduce the risk of all-cause >24-hour hospitalisation or death compared with placebo in high-risk COVID-19 patients.^13^ The oral antiviral nirmatrelvir/ritonavir inhibits the main protease of SARS-CoV-2 and has been shown to reduce the risk of progression to severe COVID-19 compared with placebo among symptomatic patients.^14,15^ Another oral antiviral, molnupiravir, a ribonucleoside prodrug with antiviral activity against SARS-CoV-2, has proven effective in reducing the risk of hospitalisation or death among at-risk, unvaccinated adults with COVID-19.^16^ Molnupiravir was first approved in November 2021 by the UK Medicines and Healthcare products Regulatory Agency for use in patients with mild-to-moderate COVID-19 deemed at risk of developing severe illness.^17^ Sotrovimab and nirmatrelvir/ritonavir received conditional marketing authorisation in December 2021 for use in patients with COVID-19 who do not require supplemental oxygen and are deemed to be at increased risk of progression to severe COVID-19.^18,19^ During the study period, the National Health Service (NHS) England clinical guidelines recommended sotrovimab and nirmatrelvir/ritonavir as first-line treatment options, and molnupiravir as a third-line option.^8^

In March 2022, the Omicron BA.2 subvariant became dominant globally.^20^ *In vitro* neutralisation assays reported a moderate fold-shift in activity for sotrovimab with Omicron BA.2 relative to wild-type SARS-CoV-2 (Wuhan-Hu-1; 16-fold).^21^ Similar fold-shifts have also been reported for other Omicron subvariants, including BA.5, which predominated in the UK from July to October 2022.^21,22^ Uncertainty remains regarding how *in vitro* antibody neutralisation activity translates to clinical effectiveness, especially for dual-action antibodies like sotrovimab, which have potent effector function, including antibody-dependent cellular cytotoxicity and antibody-dependent cellular phagocytosis.^12,23,24^ The rapid rate of SARS-CoV-2 mutation makes it challenging to assess the effectiveness of interventions against emerging variants of concern, especially when confounded by population immunity from vaccination or natural infection, which may lower the mAb titres needed for neutralisation in humans. As such, real-world evidence obtained from routine clinical practice has become increasingly important to assess treatment effectiveness outside of preclinical studies or clinical trials.

Here we describe the real-world use of, and outcomes from, early, mild-to-moderate COVID-19 treatments for the management of highest-risk patients in north-west London (NWL) following the emergence of Omicron subvariants.

## Methods

### Study design and data source

This retrospective cohort study used data from the Discover dataset, one of Europe’s largest linked longitudinal datasets.^25^ It holds depersonalised coded primary and secondary care data (starting from 1990 and 2015, respectively) for over 2.5 million patients who live and are registered with a general practitioner (GP) in NWL, a region inhabited by a highly ethnically diverse population. The data also contain information on acute health, mental health, community health and social care for these patients. The dataset is fed by data from over 400 provider organisations, including over 350 GP practices, two mental health and two community trusts, and all acute providers attended by NWL patients.

The age profile of the population in NWL is comparable to the age profile of patients across the whole of London. Furthermore, the Discover population has comparable age–sex distribution and prevalence of comorbidities to the overall UK population, but is more ethnically diverse.^25^ The dataset is accessible via Discover-NOW Health Data Research Hub for Real World Evidence through their data scientist specialists and Information Governance (IG) committee-approved analysts, hosted by Imperial College Health Partners.

The index date was defined as the earliest date of COVID-19 diagnosis or a positive polymerase chain reaction (PCR) or lateral flow test for SARS-CoV-2, where that event occurred between 1 December 2021 and 31 May 2022; if the date of the diagnosis was not available, date of treatment was used as a proxy. During the course of this study, the Omicron BA.5 variant began circulating. Given the importance of generating evidence on emerging COVID-19 strains, a *post-hoc* analysis was undertaken among patients diagnosed or treated between 1 June 2022 and 31 July 2022, a period during which Omicron BA.5 was the predominant strain in England.^26^ The baseline period was defined as the 365 days prior to index. Patient outcomes were reported during the acute period, defined as the 28-day period starting on the index date.

### Study population

Non-hospitalised patients were eligible for inclusion if they were aged ≥12 years on the index date; had a COVID-19 diagnosis/positive SARS-CoV-2 PCR or lateral flow test; and received outpatient treatment with sotrovimab, nirmatrelvir/ritonavir or molnupiravir, or received no treatment and met at least one of the NHS highest-risk criteria for receiving early treatment with sotrovimab, nirmatrelvir/ritonavir, molnupiravir or remdesivir. At the time of study, these criteria included Down’s syndrome, solid cancer, haematological diseases (including cancers), advanced renal disease, advanced liver disease, immune-mediated inflammatory disorders, immune deficiencies, HIV/AIDS, solid organ and stem cell transplant recipients and rare neurological conditions.^7^ Patients meeting the highest-risk criteria were identified via the presence of International Classification of Disease version 10 (ICD-10) and Systematized Nomenclature of Medicine (SNOMED) codes appearing at any time in the patient’s records since first registration in NWL (source datasets included in Table S1; conditions included in Table 1).

**Table 1.**
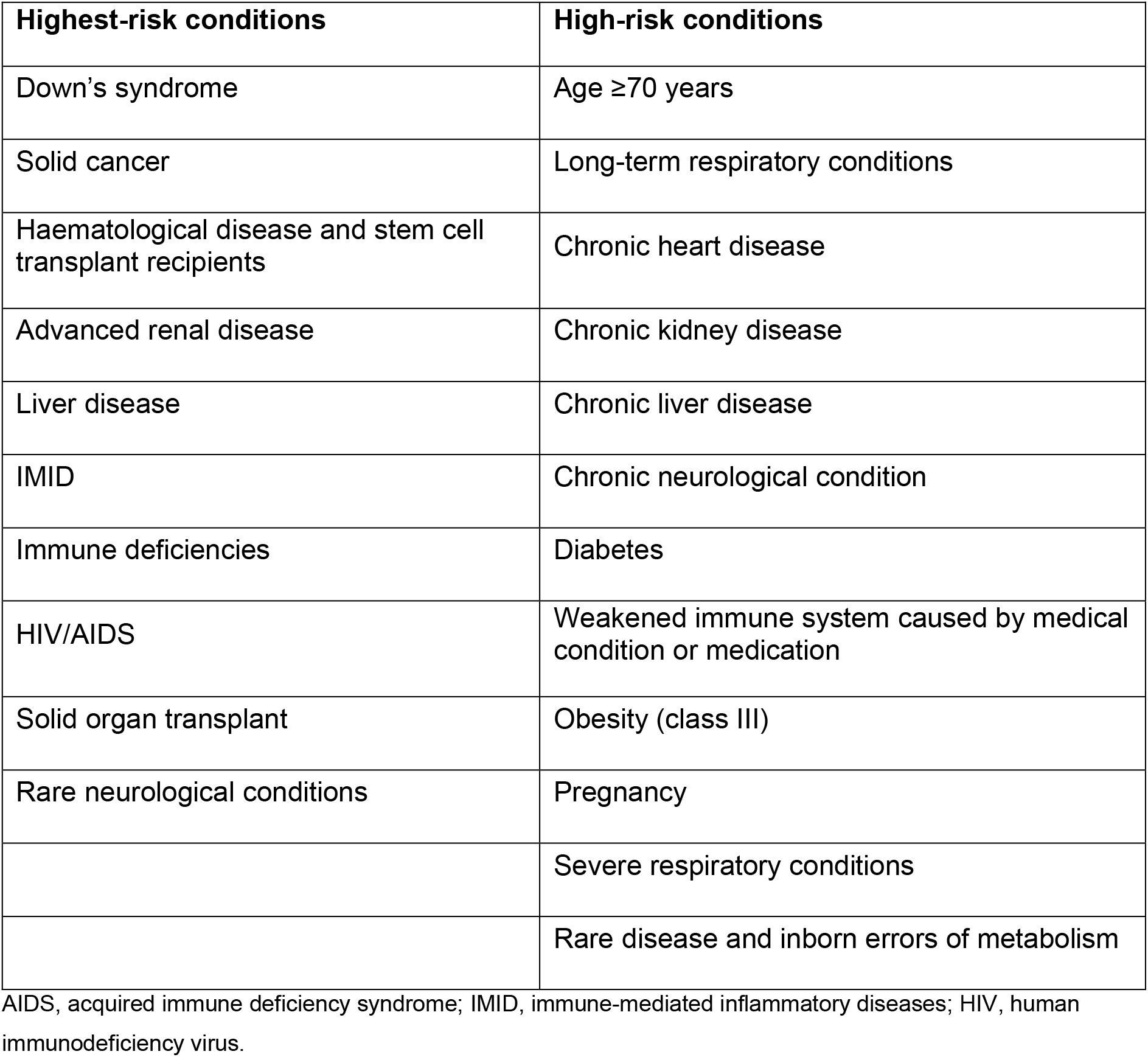
High- and highest-risk conditions criteria

Patients were excluded if they received more than one treatment (sotrovimab, nirmatrelvir/ritonavir, molnupiravir or remdesivir) in an outpatient setting during the acute period; they received remdesivir as an early treatment (due to low number of patients receiving the drug); or they received sotrovimab, nirmatrelvir/ritonavir or molnupiravir while in an inpatient setting (involving an overnight or longer stay in a hospital), or more than 28 days following first diagnosis of COVID-19.

### Study outcomes

The primary outcomes of this study were acute period clinical outcomes, namely COVID-19-related and all-cause hospitalisations. COVID-19-related hospitalisations were defined as any non-elective hospital visit for which COVID-19 (ICD-10 U07.1) was listed in the primary diagnosis field. All-cause hospitalisations were defined as non-elective hospital visits for any diagnosis. The secondary outcome reported in this study was the number of deaths due to any cause.

Patient characteristics were also recorded, including age, gender, ethnicity, vaccination status and comorbidity history. Cohorts were described in relation to ‘highest-risk’ conditions which made patients eligible for early treatment with mAbs and antiviral therapies, as mentioned above. Additionally, the cohorts were described in relation to other ‘high-risk’ conditions which may predispose patients to severe COVID-19 outcomes (Table 1).

Outcomes were reported for the following cohorts: Cohort 1) patients receiving early treatment with sotrovimab; Cohort 2) patients receiving early treatment with nirmatrelvir/ritonavir; Cohort 3) patients receiving early treatment with molnupiravir; and Cohort 4) patients at highest risk who received no early treatment.

Subgroup analyses were also undertaken. We examined highest-risk patients with advanced renal disease (defined as patients with chronic kidney disease stage 4 or 5 [including codes for peritoneal and haemodialysis] and kidney transplant recipients, or non-transplant recipients receiving a comparable level of immunosuppression; chronic kidney disease stage 1–3 reported separately as a high-risk condition). A subgroup analysis was also undertaken by age group for patients aged 18–64 and ≥65 years. We also investigated outcomes among patients diagnosed or treated during periods of Omicron BA.1 (1 December 2021–28 February 2022) and Omicron BA.2 (1 March–31 May 2022) subvariant predominance.

During our Omicron BA.1 period, 71.2% of sequenced cases across England were BA.1, while during our Omicron BA.2 period, 90.1% of sequenced cases across England were BA.2.^27^ In addition, a *post-hoc* exploratory analysis was conducted to examine patients diagnosed and treated during early BA.5 variant predominance (1 June–31 July 2022) once more recent data had become available. During this time, 70.6% of sequenced cases were attributable to the Omicron BA.5 variant.^26^

### Data analysis

Continuous variables (e.g. age) were summarised using mean, standard deviation, median, interquartile range and range. Categorical variables (e.g. gender) were described using frequencies and percentages. For clinical outcomes, 95% confidence intervals (CIs) were calculated. No inferential statistics were performed as part of the analysis. Due to information governance and data suppression rules used in the study, counts of between one and four were supressed and are reported as n<5 throughout.

## Results

### Patient demographics and baseline characteristics

The analysis included 696 patients treated with sotrovimab, 337 patients treated with nirmatrelvir/ritonavir, 470 patients treated with molnupiravir and 4,044 eligible highest-risk untreated patients (Table 2).

**Table 2.** Patient characteristics

|  | **Sotrovimab**<br>**(n=696)** | **Nirmatrelvir/**<br>**ritonavir**<br>**(n=337)** | **Molnupiravir**<br>**(n=470)** | **Untreated**<br>**(n=4,044)** |
| --- | --- | --- | --- | --- |
| **Age** |  |  |  |  |
| Mean (SD) | 58.0 (16.4) | 52.6 (15.5) | 53.7 (17.4) | 52.4 (17.5) |
| Median (IQR) | 58 (25) | 54 (22) | 54 (27) | 52 (24) |
| **Female sex, n (%)** | 356 (51.1) | 159 (47.2) | 217 (46.2) | 1,834 (45.4) |
| **Ethnicity, n (%)** |  |  |  |  |
| White | 391 (56.2) | 227 (67.4) | 261 (55.5) | 1,986 (49.1) |
| Asian/Asian British | 166 (23.9) | 45 (13.4) | 130 (27.7) | 940 (23.2) |
| Black/Black British | 60 (8.5) | 38 (11.3) | 30 (6.4) | 651 (16.1) |
| Mixed | 21 (3.0) | 8 (2.4) | 14 (3.0) | 150 (3.7) |
| Other | 45 (6.5) | 15 (4.5) | 26 (5.5) | 232 (5.7) |
| Unknown | 13 (1.9) | <5 (0.3–1.2)<sup>a</sup> | 9 (1.9) | 85 (2.1) |
| **Vaccination, n (%)** |  |  |  |  |
| No doses | 34 (4.9) | 31 (9.2) | 16 (3.4) | 460 (11.4) |
| 1 dose | 11 (1.6) | 5 (1.5) | 6 (1.3) | 96 (2.4) |
| 2 doses | 53 (7.6) | 22 (6.5) | 42 (8.9) | 698 (17.3) |
| 1 booster | 360 (51.7) | 177 (52.5) | 328 (69.8) | 2,237 (55.3) |
| >1 booster | 238 (34.2) | 102 (30.3) | 78 (16.6) | 553 (13.7) |
| **Time since last**<br>**vaccination (days)** |  |  |  |  |
| Mean (SD) | 115.5 (73.0) | 121.3 (72.7) | 88.1 (61.5) | 116.7 (78.1) |
| Median (IQR) | 106 (87) | 122 (92) | 79 (52) | 99 (98) |
| **Time to treatment**<br>**(days)<sup>b</sup>** |  |  |  |  |
| Mean (SD) | 1.7 (1.6) | 0.7 (1.6) | 1.1 (1.4) | - |
| Median (IQR) | 1 (1) | 0 (1) | 1 (1) | - |
| **Highest-risk conditions, n (%)** |  |  |  |  |
| Down's syndrome | <5 (0.1–0.6)<sup>a</sup> | 5 (1.5) | 6 (1.3) | 61 (1.5) |
| Solid cancer | 35 (5.0) | 9 (2.7) | 9 (1.9) | 380 (9.4) |
| Haematological<br>disease and stem cell<br>transplant recipients | 48 (6.9) | 11 (3.3) | 21 (4.5) | 330 (8.2) |
| Advanced renal<br>disease | 204 (29.3) | <5 (0.3–1.2)<sup>a</sup> | 60 (12.8) | 880 (21.8) |
| Liver disease | 29 (4.2) | 5 (1.5) | 22 (4.7) | 413 (10.2) |
| IMiD | 30 (4.3) | 9 (2.7) | 45 (9.6) | 415 (10.3) |
| Immune deficiencies | 50 (7.2) | 96 (28.5) | 47 (10.0) | 1,080 (26.7) |
| HIV/AIDS | 44 (6.3) | 102 (30.3) | 46 (9.8) | 1,000 (24.7) |
| Solid organ transplant | 50 (7.2) | 0 (0.0) | 32 (6.8) | 203 (5.0) |
| Rare neurological<br>conditions | 56 (8.0) | 52 (15.4) | 48 (10.2) | 438 (10.8) |
| **Number of highest-risk conditions, n (%)** |  |  |  |  |
| 0 | 273 (39.2) | 141 (41.8) | 215 (45.7) | 0 (0.0) |
| 1 | 314 (45.1) | 106 (31.5) | 175 (37.2) | 2,935 (72.6) |
| 2 | 109 (15.7) | 90 (26.7) | 80 (17.0) | 1,109 (27.4) |
| **High-risk conditions, n (%)** |  |  |  |  |
| Age ≥70 years | 190 (27.3) | 46 (13.6) | 97 (20.6) | 720 (17.8) |
| Long-term respiratory conditions | 178 (25.6) | 72 (21.4) | 108 (23.0) | 884 (21.9) |
| Chronic heart disease | 355 (51.0) | 60 (17.8) | 190 (40.4) | 1,419 (35.1) |
| Chronic kidney disease | 142 (20.4) | 18 (5.3) | 54 (11.5) | 605 (15.0) |
| Chronic liver disease | 49 (7.0) | 10 (3.0) | 30 (6.4) | 490 (12.1) |
| Chronic neurological condition | 361 (51.9) | 116 (34.4) | 201 (42.8) | 1,659 (41.0) |
| Diabetes | 176 (25.3) | 29 (8.6) | 81 (17.2) | 824 (20.4) |
| Weakened immune system caused by medical condition or medication | 122 (17.5) | 25 (7.4) | 91 (19.4) | 600 (14.8) |
| Obesity (class III) | 16 (2.3) | 5 (1.5) | 14 (3.0) | 72 (1.8) |
| Pregnancy | <5 (0.1–0.6)<sup>a</sup> | 9 (2.7) | 9 (1.9) | 224 (5.5) |
| Severe respiratory conditions | 148 (21.3) | 47 (13.9) | 84 (17.9) | 625 (15.5) |
| Rare disease and inborn errors of metabolism | <5 (0.1–0.6)<sup>a</sup> | 9 (2.7) | 9 (1.9) | 224 (5.5) |
| **Number of high-risk conditions, n (%)** |  |  |  |  |
| 0 | 82 (11.8) | 105 (31.2) | 69 (14.7) | 856 (21.2) |
| 1 | 144 (20.7) | 112 (33.2) | 139 (29.6) | 968 (23.9) |
| 2 | 139 (20.0) | 62 (18.4) | 97 (20.6) | 798 (19.7) |
| 3 | 140 (20.1) | 36 (10.7) | 78 (16.6) | 585 (14.5) |
| 4 | 99 (14.2) | 13 (3.9) | 46 (9.8) | 407 (10.1) |
| 5+ | 92 (13.2) | 9 (2.7) | 41 (8.7) | 430 (10.6) |

<sup>a</sup>Due to a total sample size of <5, this number was suppressed; a range of 1–4 events has been used to calculate the range given; <sup>b</sup>No COVID-19 diagnosis date was available for 52 patients on sotrovimab, 121 patients on nirmatrlvir/ritonavir and 16 patients on molnupiravir. A time to treatment of 0 days was assumed for these patients. AIDS, acquired immune deficiency syndrome; HIV, human immunodeficiency virus; IMID, immune-mediated inflammatory disease; IQR, interquartile range; SD, standard deviation.

A high percentage of patients treated with sotrovimab had advanced renal disease (29.3%, n=204/696), while lower percentages were reported for other highest-risk comorbidities (solid organ transplant recipients: 7.2% [n=50/696]; rare neurological conditions: 8.0% [n=56/696]; haematological disease or stem cell transplant recipient: 6.9% [n=48/696]). Of the patients identified to receive nirmatrelvir/ritonavir, 28.5% (n=96/337) had immune deficiencies, 30.3% (n=102/337) had HIV/AIDS and 15.4% (n=52/337) had rare neurological disorders. Advanced renal disease was the most frequently reported highest-risk condition for patients treated with molnupiravir (12.8%, n=60/470).

Two or more highest-risk conditions were recorded for 15.7% (n=109/696) of patients treated with sotrovimab, 26.7% (n=90/337) of patients treated with nirmatrelvir/ritonavir, 17.0% (n=80/470) of those treated with molnupiravir and 27.4% (n=1,109/4,044) of untreated patients. No highest-risk conditions were recorded in 39.2% (n=273/696) of patients treated with sotrovimab, 41.8% (n=141/337) of patients treated with nirmatrelvir/ritonavir and 45.7% (n=215/470) of patients treated with molnupiravir. No untreated patients had zero highest-risk conditions since inclusion within this cohort was based on the presence of an identifiable highest-risk condition.

A high percentage of sotrovimab-treated patients had high-risk comorbidities such as chronic heart disease (51.0%, n=355/696), chronic kidney disease (20.4%, n=142/696), chronic neurological conditions (51.9%, n=361/696), diabetes (25.3%, n=176/696) and severe respiratory conditions (21.3%, n=148/696). For patients treated with nirmatrelvir/ritonavir, 17.8% (n=60/337) had chronic heart disease, 5.3% (n=18/337) had chronic kidney disease, 34.4% (n=116/337) had chronic neurological conditions, 8.6% (n=29/337) had diabetes and 13.9% (n=47/337) had severe respiratory conditions. Patients treated with molnupiravir also presented with high-risk comorbidities: 40.4% (n=190/470) had chronic heart disease, 11.5% (n=54/470) had chronic kidney disease, 42.8% (n=201/470) had chronic neurological conditions, 17.2% (n=81/470) presented with diabetes and 17.9% (n=84/470) had severe respiratory conditions. Similarly, 35.1% (n=1,419/4,044) of untreated patients had chronic heart disease, 15.0% (n=605/4,044) had chronic kidney disease, 41.0% (n=1,659/4,044) had chronic neurological conditions, 20.4% (n=824/4,044) had diabetes and 15.5% (n=625/4,044) presented with severe respiratory conditions.

The proportion of patients with at least three high-risk comorbidities was 47.6% (n=331/696) among those treated with sotrovimab, 17.2% (n=58/337) among those treated with nirmatrelvir/ritonavir, 35.1% (n=165/470) for those treated with molnupiravir and 35.2% (n=1,422/4,044) for patients who were untreated. Percentages for patients with no or unidentifiable high-risk conditions were 11.8% for sotrovimab (n=82/696), 14.7% for molnupiravir (n=69/470), 31.2% for nirmatrelvir/ritonavir (n=105/337) and 21.2% for untreated patients (n=856/4,044).

In addition, patients aged ≥70 years accounted for 27.3% (n=190/696) of the sotrovimab-treated group, 13.6% (n=46/337) of those treated with nirmatrelvir/ritonavir, 20.6% (n=97/470) of those treated with molnupiravir and 17.8% (n=720/4,044) of untreated patients. A high proportion of untreated patients were of Black/Black British ethnicity (16.1%, n=651/4,044); this was 8.6% (n=60/696) for sotrovimab-, 11.3% (n=38/337) for nirmatrelvir/ritonavir- and 6.4% (n=30/470) for molnupiravir-treated patients. A low proportion of untreated patients had received more than one booster vaccination (13.7%, n=553/4,044). The proportions of patients treated with sotrovimab, nirmatrelvir/ritonavir and molnupiravir who received more than one booster were 34.2% (n=238/696), 30.3% (n=102/337) and 16.6% (n=78/470), respectively.

#### Acute period clinical outcomes

A low proportion of patients treated with sotrovimab (n=5/696, 0.7% [95% CI 0.1%, 1.3%]), nirmatrelvir/ritonavir (n<5/337, 0.3–1.2%) or molnupiravir (n=10/470, 2.1% [95% CI 0.8%, 3.4%]) experienced COVID-19-related hospitalisations. For untreated patients, the percentage of COVID-19-related hospitalisations was 2.8% (95% CI 2.3%, 3.3%; n=114/4,044) (Table 3). The percentage of patients who experienced an all-cause hospitalisation was 5.0% (95% CI 3.4%, 6.6%; n=35/696) for sotrovimab-treated patients, 1.5% (95% CI 0.2%, 2.8%; n=5/337) for nirmatrelvir/ritonavir, 4.0% (95% CI 2.2%, 5.8%; n=19/470) for molnupiravir and 6.2% (95% CI 5.5%, 6.9%; n=251/4,044) for untreated patients. The proportion of patients dying within 28 days of the index period was similarly low across all cohorts (<2% in each).

**Table 3.** Acute period clinical outcomes

|  | **Sotrovimab**<br>**(n=696)** | **Nirmatrelvir/**<br>**ritonavir**<br>**(n=337)** | **Molnupiravir**<br>**(n=470)** | **Untreated**<br>**(n=4,044)** |
| --- | --- | --- | --- | --- |
| COVID-19-related hospitalisation, n (%) | 5 (0.7) | <5 (0.3–1.2<sup>a</sup>) | 10 (2.1) | 114 (2.8) |
| 95% CI | 0.1, 1.3 | - | 0.8, 3.4 | 2.3, 3.3 |
| All-cause hospitalisation, n (%) | 35 (5.0) | 5 (1.5) | 19 (4.0) | 251 (6.2) |
| 95% CI | 3.4, 6.6 | 0.2, 2.8 | 2.2, 5.8 | 5.5, 6.9 |
| Patient death within 1 month of index, n (%) | 8 (1.1) | <5 (0.3–1.2<sup>a</sup>) | 7 (1.5) | 75 (1.9) |
| 95% CI | 0.3, 1.9 | - | 0.4, 2.6 | 1.5, 2.3 |

<sup>a</sup>Due to a total sample size of <5, this number was suppressed; a range of 1–4 events has been used to calculate the range given. CI, confidence interval.

### Characteristics and outcomes of patients with advanced renal disease, untreated or treated with sotrovimab

Due to the low numbers of patients with advanced renal disease being treated with nirmatrelvir/ritonavir (n<5) or molnupiravir (n=60), characteristics (Table 4) and acute-period clinical outcomes (Table 5) have only been reported for the sotrovimab-treated and untreated patient cohorts. In total, 204 patients treated with sotrovimab and 880 untreated patients were included in this subgroup analysis.

**Table 4.**
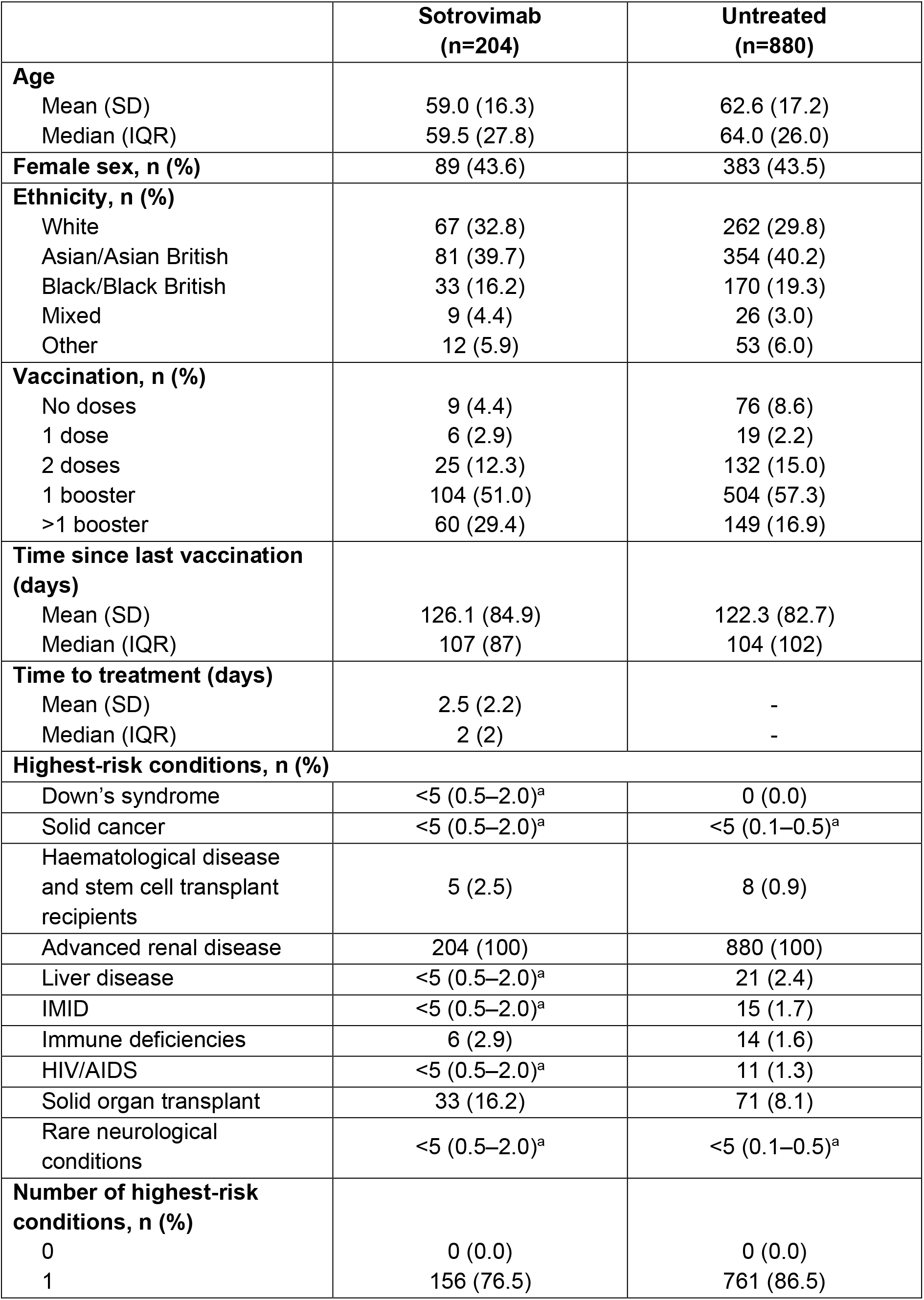
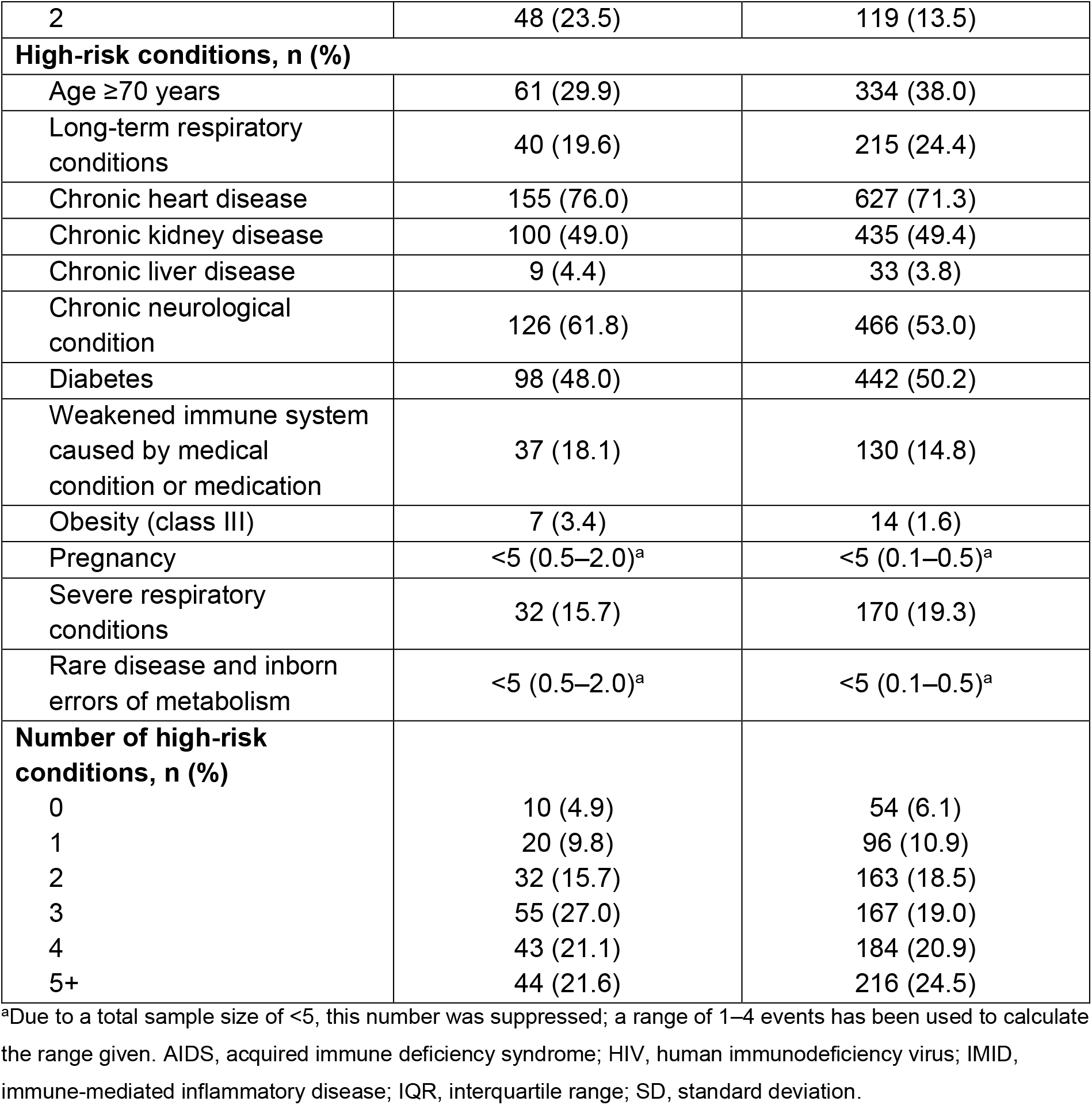
Characteristics of patients with advanced renal disease, untreated or treated with sotrovimab

**Table 5.** Acute period clinical outcomes amongst patients with renal disease

|  | **Sotrovimab**<br>**(n=204)** | **Untreated**<br>**(n=880)** |
| --- | --- | --- |
| COVID-19-related hospitalisation, n (%) | <5 (0.5–2.0<sup>a</sup>) | 39 (4.4) |
| 95% CI | - | 3.0, 5.8 |
| All-cause hospitalisation, n (%) | 17 (8.3) | 94 (10.7) |
| 95% CI | 4.5, 12.1 | 8.7, 12.7 |
| Patient death within 1 month of index, n (%) | <5 (0.5–2.0<sup>a</sup>) | 36 (4.1) |
| 95% CI | - | 2.8, 5.4 |

<sup>a</sup>Due to a total sample size of <5, this number was suppressed; a range of 1–4 events has been used to calculate the range given. CI, confidence interval.

Amongst patients with advanced renal disease, the characteristics of those treated with sotrovimab were generally similar to untreated patients. The proportion of patients who received more than one booster vaccine was 29.4% (n=60/204) for sotrovimab-treated patients and 16.9% (n=149/880) for untreated patients. The percentage of patients who had a solid organ transplant was 16.2% (n=33/204) for patients who were treated with sotrovimab, and 8.1% (n=71/880) for untreated patients. In total, 29.9% (n=61/204) of sotrovimab-treated patients and 38.0% (n=334/880) of untreated patients with advanced renal disease were aged ≥70 years.

Fewer than five of 204 (0.5–2.0%) patients treated with sotrovimab, and 39 of 880 (4.4% [95% CI 3.0%, 5.8%]) untreated patients experienced a COVID-19-related hospitalisation during the acute period (Table 5). Similarly, fewer than five (0.5–2.0%) patients treated with sotrovimab and 36 (4.1% [95% CI 2.8%, 5.4%]) untreated patients died within 1 month of index. All-cause hospitalisation was experienced by 8.3% (95% CI 4.5%, 12.1%; n=17/204) of sotrovimab-treated patients and 10.7% (95% CI 8.7%, 12.7%; n=94/880) of untreated patients.

### Characteristics and outcomes of patients aged 18–64 and ≥65 years

Characteristics among patients aged 18–64 years (n=439 for sotrovimab; n=263 for nirmatrelvir/ritonavir; n=339 for molnupiravir; n=2,977 for untreated patients) were similar to those of patients aged ≥65 years (n=257 for sotrovimab; n=73 for nirmatrelvir/ritonavir; n=131 for molnupiravir; n=999 for untreated patients) (Tables 6 and 7). It should be noted that due to overall small patient numbers, those aged 12–17 years were not included in the age subgroup analysis.

**Table 6.** Characteristics of patients aged 18–64 years

|  | **Sotrovimab**<br>**(n=439)** | **Nirmatrelvir/**<br>**ritonavir**<br>**(n=263)** | **Molnupiravir**<br>**(n=339)** | **Untreated**<br>**(n=2,977)** |
| --- | --- | --- | --- | --- |
| **Age** |  |  |  |  |
| Mean (SD) | 48.0 (11.2) | 46.9 (11.9) | 45.3 (11.9) | 45.5 (11.8) |
| Median (IQR) | 50 (17) | 49 (19) | 47 (19) | 46 (19) |
| **Female sex, n (%)** | 230 (52.4) | 132 (50.2) | 155 (45.7) | 1,325 (44.5) |
| **Ethnicity, n (%)** |  |  |  |  |
| White | 240 (54.7) | 169 (64.3) | 177 (52.2) | 1,430 (48.0) |
| Asian/Asian British | 104 (23.7) | 37 (14.1) | 99 (29.2) | 645 (21.7) |
| Black/Black British | 44 (10.0) | 37 (14.1) | 23 (6.8) | 533 (17.9) |
| Mixed | 16 (3.6) | 6 (2.3) | 12 (3.5) | 125 (4.2) |
| Other | 29 (6.6) | 10 (3.8) | 20 (5.9) | 179 (6.0) |
| Unknown | 6 (1.4) | <5 (0.4–1.5)<sup>a</sup> | 8 (2.4) | 65 (2.2) |
| **Vaccination, n (%)** |  |  |  |  |
| No doses | 27 (6.2) | 28 (10.6) | 13 (3.8) | 370 (12.4) |
| 1 dose | 6 (1.4) | 6 (2.3) | 5 (1.5) | 67 (2.3) |
| 2 doses | 42 (9.6) | 18 (6.8) | 38 (11.2) | 586 (19.7) |
| 1 booster | 233 (53.1) | 146 (55.5) | 231 (68.1) | 1,589 (53.4) |
| >1 booster | 131 (29.8) | 65 (24.7) | 52 (15.3) | 365 (12.3) |
| **Time since last**<br>**vaccination (days)** |  |  |  |  |
| Mean (SD) | 118.0 (72.4) | 122.0 (71.1) | 88.0 (65.8) | 117.4 (78.7) |
| Median (IQR) | 106 (83.0) | 121.5 (92.3) | 75 (53.8) | 99 (103.0) |
| **Time to treatment**<br>**(days)** |  |  |  |  |
| Mean (SD) | 1.5 (1.5) | 0.7 (1.7) | 1.0 (1.1) | - |
| Median (IQR) | 1 (1) | 0 (1) | 1 (1) | - |
| **Highest-risk conditions, n (%)** |  |  |  |  |
| Down's syndrome | <5 (0.2–0.9)<sup>a</sup> | 5 (1.9) | 5 (1.5) | 39 (1.3) |
| Solid cancer | 14 (3.2) | 7 (2.7) | 6 (1.8) | 233 (7.8) |
| Haematological<br>disease and stem<br>cell transplant<br>recipients | 23 (5.2) | 10 (3.8) | 15 (4.4) | 244 (8.2) |
| Advanced renal<br>disease | 120 (27.3) | 0 (0.0) | 42 (12.4) | 462 (15.5) |
| Liver disease | 17 (3.9) | 5 (1.9) | 14 (4.1) | 249 (8.4) |
| IMiD | 29 (6.6) | 8 (3.0) | 40 (11.8) | 352 (11.8) |
| Immune<br>deficiencies | 42 (9.6) | 76 (28.9) | 43 (12.7) | 984 (33.1) |
| HIV/AIDS | 37 (8.4) | 82 (31.2) | 44 (13.0) | 939 (31.5) |
| Solid organ<br>transplant | 35 (8.0) | 0 (0.0) | 25 (7.4) | 153 (5.1) |
| Rare neurological conditions | 43 (9.8) | 45 (17.1) | 39 (11.5) | 352 (11.8) |
| **Number of highest-risk conditions, n (%)** |  |  |  |  |
| 0 | 166 (37.8) | 100 (38.0) | 132 (38.9) | 0 (0.0) |
| 1 | 195 (44.4) | 91 (34.6) | 141 (41.6) | 1,986 (66.7) |
| 2 | 78 (17.8) | 72 (27.4) | 66 (19.5) | 991 (33.3) |
| **High-risk conditions, n (%)** |  |  |  |  |
| Long-term respiratory conditions | 105 (23.9) | 60 (22.8) | 67 (19.8) | 600 (20.2) |
| Chronic heart disease | 169 (38.5) | 38 (14.4) | 113 (33.3) | 739 (24.8) |
| Chronic kidney disease | 60 (13.7) | 6 (2.3) | 21 (6.2) | 258 (8.7) |
| Chronic liver disease | 33 (7.5) | 9 (3.4) | 22 (6.5) | 311 (10.4) |
| Chronic neurological condition | 207 (47.2) | 87 (33.1) | 146 (43.1) | 1,006 (33.8) |
| Diabetes | 80 (18.2) | 11 (4.2) | 45 (13.3) | 392 (13.2) |
| Weakened immune system caused by medical condition or medication | 96 (21.9) | 20 (7.6) | 80 (23.6) | 495 (16.6) |
| Obesity (class III) | 8 (1.8) | 5 (1.9) | 12 (3.5) | 58 (1.9) |
| Pregnancy | <5 (0.2–0.9)<sup>a</sup> | 9 (3.4) | 8 (2.4) | 186 (6.2) |
| Severe respiratory conditions | 83 (18.9) | 38 (14.4) | 51 (15.0) | 400 (13.4) |
| Rare disease and inborn errors of metabolism | <5 (0.2–0.9)<sup>a</sup> | 9 (3.4) | 8 (2.4) | 186 (6.2) |
| **Number of high-risk conditions, n (%)** |  |  |  |  |
| 0 | 73 (16.6) | 97 (36.9) | 67 (19.8) | 822 (27.6) |
| 1 | 122 (27.8) | 89 (33.8) | 116 (34.2) | 861 (28.9) |
| 2 | 99 (22.6) | 43 (16.3) | 64 (18.9) | 621 (20.9) |
| 3 | 80 (18.2) | 23 (8.7) | 53 (15.6) | 352 (11.8) |
| 4 | 44 (10.0) | 9 (3.4) | 26 (7.7) | 192 (6.4) |
| 5+ | 21 (4.8) | <5 (0.4–1.5)<sup>a</sup> | 13 (3.8) | 129 (4.3) |

<sup>a</sup>Due to a total sample size of <5, this number was suppressed; a range of 1–4 events has been used to calculate the range given. AIDS, acquired immune deficiency syndrome; HIV, human immunodeficiency virus; IMID, immune-mediated inflammatory disease; IQR, interquartile range; SD, standard deviation.

**Table 7.** Characteristics of patients aged ≥65 years

|  | **Sotrovimab**<br>**(n=257)** | **Nirmatrelvir/**<br>**ritonavir**<br>**(n=73)** | **Molnupiravir**<br>**(n=131)** | **Untreated**<br>**(n=999)** |
| --- | --- | --- | --- | --- |
| **Age** |  |  |  |  |
| Mean (SD) | 75.1 (7.3) | 73.2 (6.5) | 75.5 (7.8) | 75.3 (7.7) |
| Median (IQR) | 74 (11) | 73 (10) | 75 (11) | 74 (12) |
| **Female sex, n (%)** | 126 (49.0) | 26 (35.6) | 62 (47.3) | 477 (47.7) |
| **Ethnicity, n (%)** |  |  |  |  |
| White | 151 (58.8) | 57 (78.1) | 84 (64.1) | 539 (54.0) |
| Asian/Asian British | 62 (24.1) | 8 (11.0) | 31 (23.7) | 276 (27.6) |
| Black/Black British | 16 (6.2) | <5 (1.4–5.5)<sup>a</sup> | 7 (5.3) | 100 (10.0) |
| Mixed | 5 (1.9) | <5 (1.4–5.5)<sup>a</sup> | <5 (0.8–3.1)<sup>a</sup> | 23 (2.3) |
| Other | 16 (6.2) | 5 (6.8) | 6 (4.6) | 44 (4.4) |
| Unknown | 7 (2.7) | 0 (0.0) | <5 (0.8–3.1)<sup>a</sup> | 17 (1.7) |
| **Vaccination, n (%)** |  |  |  |  |
| No doses | 7 (2.7) | <5 (1.4–5.5)<sup>a</sup> | <5 (0.8–3.1)<sup>a</sup> | 45 (4.5) |
| 1 dose | 5 (1.9) | 0 (0.0) | <5 (0.8–3.1)<sup>a</sup> | 12 (1.2) |
| 2 doses | 11 (4.3) | <5 (1.4–5.5)<sup>a</sup> | 5 (3.8) | 108 (10.8) |
| 1 booster | 127 (49.4) | 31 (42.5) | 97 (74.0) | 646 (64.7) |
| >1 booster | 107 (41.6) | 36 (49.3) | 26 (19.8) | 188 (18.8) |
| **Time since last**<br>**vaccination (days)** |  |  |  |  |
| Mean (SD) | 111.4 (74.0) | 117.6 (78.0) | 88.2 (49.3) | 116.1 (76.8) |
| Median (IQR) | 104 (93) | 120 (92) | 83 (33) | 101 (91) |
| **Time to treatment**<br>**(days)** |  |  |  |  |
| Mean (SD) | 1.9 (1.8) | 0.7 (1.0) | 1.3 (2.0) | - |
| Median (IQR) | 1 (2) | 0 (1) | 1 (2) | - |
| **Highest-risk conditions, n (%)** |  |  |  |  |
| Down's syndrome | 0 (0.0) | 0 (0.0) | <5 (0.8–3.1)<sup>a</sup> | <5 (0.1–0.4)<sup>a</sup> |
| Solid cancer | 21 (8.2) | <5 (1.4–5.5)<sup>a</sup> | <5 (0.8–3.1)<sup>a</sup> | 146 (14.6) |
| Haematological<br>disease and stem<br>cell transplant<br>recipients | 25 (9.7) | <5 (1.4–5.5)<sup>a</sup> | 6 (4.6) | 63 (6.3) |
| Advanced renal<br>disease | 84 (32.7) | <5 (1.4–5.5)<sup>a</sup> | 18 (13.7) | 417 (41.7) |
| Liver disease | 12 (4.7) | 0 (0.0) | 8 (6.1) | 162 (16.2) |
| IMiD | <5 (0.4–1.6)<sup>a</sup> | <5 (1.4–5.5)<sup>a</sup> | 5 (3.8) | 55 (5.5) |
| Immune<br>deficiencies | 8 (3.1) | 20 (27.4) | <5 (0.8–3.1)<sup>a</sup> | 84 (8.4) |
| HIV/AIDS | 7 (2.7) | 20 (27.4) | <5 (0.8–3.1)<sup>a</sup> | 61 (6.1) |
| Solid organ<br>transplant | 15 (5.8) | 0 (0.0) | 7 (5.3) | 45 (4.5) |
| Rare neurological conditions | 13 (5.1) | 7 (9.6) | 9 (6.9) | 85 (8.5) |
| **Number of highest-risk conditions, n (%)** |  |  |  |  |
| 0 | 107 (41.6) | 40 (54.8) | 83 (63.4) | 0 (0.0) |
| 1 | 119 (46.3) | 15 (20.5) | 34 (26.0) | 885 (88.6) |
| 2 | 31 (12.1) | 18 (24.7) | 14 (10.7) | 114 (11.4) |
| **High-risk conditions, n (%)** |  |  |  |  |
| Age ≥70 years | 190 (73.9) | 46 (63.0) | 97 (74.0) | 720 (72.1) |
| Long-term respiratory conditions | 73 (28.4) | 12 (16.4) | 41 (31.3) | 270 (27.0) |
| Chronic heart disease | 186 (72.4) | 21 (28.8) | 77 (58.8) | 652 (65.3) |
| Chronic kidney disease | 82 (31.9) | 12 (16.4) | 33 (25.2) | 347 (34.7) |
| Chronic liver disease | 16 (6.2) | <5 (1.4–5.5)<sup>a</sup> | 8 (6.1) | 177 (17.7) |
| Chronic neurological condition | 154 (59.9) | 28 (38.4) | 55 (42.0) | 618 (61.9) |
| Diabetes | 96 (37.4) | 18 (24.7) | 36 (27.5) | 431 (43.1) |
| Weakened immune system caused by medical condition or medication | 26 (10.1) | 5 (6.8) | 11 (8.4) | 96 (9.6) |
| Obesity (class III) | 8 (3.1) | 0 (0.0) | <5 (0.8–3.1)<sup>a</sup> | 14 (1.4) |
| Pregnancy | 0 (0.0) | 0 (0.0) | <5 (0.8–3.1)<sup>a</sup> | 15 (1.5) |
| Severe respiratory conditions | 65 (25.3) | 9 (12.3) | 33 (25.2) | 218 (21.8) |
| Rare disease and inborn errors of metabolism | 0 (0.0) | 0 (0.0) | <5 (0.8–3.1)<sup>a</sup> | 15 (1.5) |
| **Number of high-risk conditions, n (%)** |  |  |  |  |
| 0 | 9 (3.5) | 8 (11.0) | <5 (0.8–3.1)<sup>a</sup> | 28 (2.8) |
| 1 | 22 (8.6) | 23 (31.5) | 23 (17.6) | 89 (8.9) |
| 2 | 40 (15.6) | 18 (24.7) | 33 (25.2) | 156 (15.6) |
| 3 | 60 (23.3) | 13 (17.8) | 25 (19.1) | 218 (21.8) |
| 4 | 55 (21.4) | 4 (5.5) | 20 (15.3) | 211 (21.1) |
| 5+ | 71 (27.6) | 7 (9.6) | 28 (21.4) | 297 (29.7) |

<sup>a</sup>Due to a total sample size of <5, this number was suppressed; a range of 1–4 events has been used to calculate the range given. AIDS, acquired immune deficiency syndrome; HIV, human immunodeficiency virus; IMID, immune-mediated inflammatory disease; IQR, interquartile range; SD, standard deviation.

The percentages of patients aged 18–64 years who had more than one booster vaccination at the time of the study were 29.8% (n=131/439) for patients treated with sotrovimab, 24.7% (n=65/263) for those treated with nirmatrelvir/ritonavir, 15.3% (n=52/339) for patients treated with molnupiravir and 12.3% (n=365/2,977) for untreated patients.

Amongst patients aged 18–64 years, advanced renal disease was recorded in 27.3% (n=120/439) of sotrovimab-treated patients, 12.4% (n=43/339) of patients treated with molnupiravir, 15.5% (n=462/2,977) of untreated patients and none of the patients treated with nirmatrelvir/ritonavir. Immune deficiencies and HIV/AIDS were also recorded in patients aged 18–64 years: 9.6% (n=42/439) and 8.4% (n=37/439) for patients treated with sotrovimab, 28.9% (n=76/263) and 31.2% (n=82/263) for those treated with nirmatrelvir/ritonavir, 12.7% (n=43/339) and 13.0% (n=44/339) for patients treated with molnupiravir and 33.1% (n=984/2,977) and 31.5% (n=939/2,977) for untreated patients, respectively. Percentages for rare neurological conditions were 9.8% (n=43/439) for sotrovimab-treated patients, 17.1% (n=45/263) for nirmatrelvir/ritonavir, 11.5% (n=39/339) for molnupiravir and 11.8% (n=352/2,977) for untreated patients. At least two highest-risk conditions were recorded in 17.8% (n=78/439) of sotrovimab-treated patients, 27.4% (n=72/263) of patients treated with nirmatrelvir/ritonavir, 19.5% (n=66/339) of those treated with molnupiravir and 33.3% (n=991/2,977) of untreated patients. No highest-risk conditions were reported in 37.8% (n=166/439) of sotrovimab-treated patients, 38.0% (n=100/263) of nirmatrelvir/ritonavir-treated patients and 38.9% (n=132/339) of those treated with molnupiravir.

High-risk conditions such as chronic heart disease, chronic kidney disease, chronic neurological conditions or diabetes were also reported for patients aged 18–64 years. Respective percentages for these comorbidities were 38.5% (n=169/439), 13.7% (n=60/439), 47.2% (n=207/439) and 18.2% (n=80/439) for sotrovimab-treated patients, 14.4% (n=38/263), 2.3% (n=6/263), 33.1% (n=87/263) and 4.2% (n=11/263) for patients treated with nirmatrelvir/ritonavir, 33.3% (n=113/339), 6.2% (n=21/339), 43.1% (n=146/339) and 13.3% (n=45/339) for those treated with molnupiravir, and 24.8% (n=739/2,977), 8.7% (n=258/2,977), 33.8% (n=1,006/2,977) and 13.2% (n=392/2,977) for untreated patients. At least three high-risk conditions were recorded in 33.0% (n=145/439) of sotrovimab-treated patients, 12.9% (n=34/263) of patients treated with nirmatrelvir/ritonavir, 27.1% (n=92/339) of those treated with molnupiravir and 22.6% (n=673/2,977) of untreated patients.

The proportion of patients aged ≥65 years who had more than one booster vaccination at the time of the study was 41.6% (n=107/257) among those treated with sotrovimab, 49.3% (n=36/73) among those treated with nirmatrelvir/ritonavir, 19.8% (n=26/131) for those treated with molnupiravir and 18.8% (n=188/999) among untreated patients.

Highest-risk conditions were also reported for the patients aged ≥65 years, with the most common being advanced renal disease (32.7% [n=84/257] for sotrovimab-treated patients; 1.4–5.5% [n<5/73] for patients treated with nirmatrelvir/ritonavir; 13.7% [n=18/131] for those treated with molnupiravir; and 41.7% [n=417/999] for untreated patients), immune deficiencies (3.1% [n=8/257] for sotrovimab-treated patients; 27.4% [n=20/73] for patients treated with nirmatrelvir/ritonavir; 0.8–3.1% [n<5/131] for those treated with molnupiravir and 8.4% [n=84/999] for untreated patients), and rare neurological conditions (5.1% [n=13/257] for sotrovimab-treated patients; 9.6% [n=7/73] for patients treated with nirmatrelvir/ritonavir; 6.9% [n=9/131] for those treated with molnupiravir and 8.5% [n=85/999] for untreated patients). Two or more highest-risk condition were recorded for 12.1% (n=31/257) of sotrovimab-treated patients aged ≥65 years. The percentages were 24.7% (n=18/73) for nirmatrelvir/ritonavir, 10.7% (n=14/131) for molnupiravir and 11.4% (n=114/999) for untreated patients. No identifiable highest-risk conditions were reported in 41.6% (n=107/257) of sotrovimab-treated patients, 54.8% (n=40/73) of nirmatrelvir/ritonavir-treated patients and 63.4% (n=83/131) of those treated with molnupiravir.

The most frequently reported high-risk conditions for patients aged ≥65 years were age ≥70 and chronic heart disease, reported for 73.9% (n=190/257) and 72.4% (n=186/257) of patients treated with sotrovimab, 63.0% (n=46/73) and 28.8% (n=21/73) of those treated with nirmatrelvir/ritonavir, 74.0% (n=97/131) and 58.8% (n=77/131) of patients treated with molnupiravir and 72.1% (n=720/999) and 65.3% (n=652/999) of untreated patients, respectively. Similarly high percentages were recorded for chronic neurological conditions across all cohorts (59.9% [n=154/257], 38.4% [n=28/73], 42.0% [n=55/131] and 61.9% [n=618/999], respectively). Chronic kidney disease was reported for 31.9% (n=82/257), 16.4% (n=12/73), 25.2% (n=33/131) and 34.7% (n=347/999) of the cohorts, respectively, and diabetes for 37.4% (n=96/257), 24.7% (n=18/73), 27.5% (n=36/131) and 43.1% (431/999), respectively. Overall, at least three high-risk conditions were recorded in 72.4% (n=186/257) of patients treated with sotrovimab, 32.9% (n=24/73) of those treated with nirmatrelvir/ritonavir, 55.7% (n=73/131) of patients treated with molnupiravir and 72.7% (n=726/999) of untreated patients.

Table 8 details acute period clinical outcomes for those aged 18–64 years. As for the overall population, a low proportion of patients aged 18–64 years who were treated with sotrovimab (0.2–0.9%, n<5/439), nirmatrelvir/ritonavir (0.0%, n=0/263) or molnupiravir (n=8/339, 2.4% [95% CI 0.8, 4.0]) had COVID-19-related hospitalisations during the acute clinical period. This was 2.1% (n=64/2,977) for untreated patients. Similarly, the proportion of patients treated with nirmatrelvir/ritonavir who had an all-cause hospitalisation during this period was low (0.4–1.5%, n<5/263). For those treated with sotrovimab or molnupiravir or who were untreated, the proportions of patients with all-cause hospitalisations were 4.3% (95% CI 2.4%, 6.2%; n=19/439), 4.4% (95% CI 2.2%, 6.6%; n=15/339) and 4.4% (95% CI 3.7%, 5.1%; n=131/2,977), respectively. No patient deaths were recorded for nirmatrelvir/ritonavir within 1 month of index, while fewer than 5 deaths were reported for patients treated with sotrovimab (0.2–0.9%) and patients treated with molnupiravir (0.3–1.2%). A total of 17 deaths (0.6% [95% CI 0.3%, 0.9%]) were reported within 1 month of index amongst the 2,977 patients who received no treatment.

**Table 8.** Acute period clinical outcomes amongst patients aged 18–64 years

|  | **Sotrovimab**<br>**(n=439)** | **Nirmatrelvir/**<br>**ritonavir**<br>**(n=263)** | **Molnupiravir**<br>**(n=339)** | **Untreated**<br>**(n=2,977)** |
| --- | --- | --- | --- | --- |
| COVID-19-related hospitalisation, n (%) | <5 (0.2–0.9<sup>a</sup>) | 0 (0.0) | 8 (2.4) | 64 (2.1) |
| 95% CI | - | - | 0.8, 4.0 | 1.6, 2.6 |
| All-cause hospitalisation, n (%) | 19 (4.3) | <5 (0.4–1.5<sup>a</sup>) | 15 (4.4) | 131 (4.4) |
| 95% CI | 2.4, 6.2 | - | 2.2, 6.6 | 3.7, 5.1 |
| Patient death within 1 month of index, n (%) | <5 (0.2–0.9<sup>a</sup>) | 0 (0.0) | <5 (0.3–1.2<sup>a</sup>) | 17 (0.6) |
| 95% CI | - | - | - | 0.3, 0.9 |

<sup>a</sup>Due to a total sample size of <5, this number was suppressed; a range of 1–4 events has been used to calculate the range given. CI, confidence interval.

Table 9 details acute period clinical outcomes for patients aged ≥65 years. The proportion of patients treated with sotrovimab who experienced a COVID-19-related hospitalisation was 0.4–1.6% (n<5/257), and was 5.0% (95% CI 3.6%, 6.4%; n=50/999) for untreated patients. In addition, a low proportion of patients treated with sotrovimab (6.2% [95% CI 3.2%, 9.2%], n=16/257) experienced an all-cause hospitalisation during the acute clinical period. Similarly, a low proportion of patients treated with sotrovimab (0.4–1.6%, n<5/257) died within 1 month of their index date. This percentage was 5.8% (95% CI 4.3%, 7.3%; n=58/999) for untreated patients. Due to the small sample size in the subgroup aged 65 years and above, acute period clinical outcome results for patients treated with nirmatrelvir/ritonavir and molnupiravir should be treated with caution.

**Table 9.** Acute period clinical outcomes amongst patients aged ≥65 years

|  | **Sotrovimab**<br>**(n=257)** | **Nirmatrelvir/**<br>**ritonavir**<br>**(n=73)** | **Molnupiravir**<br>**(n=131)** | **Untreated**<br>**(n=999)** |
| --- | --- | --- | --- | --- |
| COVID-19-related hospitalisation, n (%) | <5 (0.4–1.6<sup>a</sup>) | <5 (1.4–5.5<sup>a</sup>) | <5 (0.8–3.1<sup>a</sup>) | 50 (5.0) |
| 95% CI | - | - | - | 3.6, 6.4 |
| All-cause hospitalisation, n (%) | 16 (6.2) | <5 (1.4–5.5<sup>a</sup>) | <5 (0.8–3.1<sup>a</sup>) | 117 (11.7) |
| 95% CI | 3.2, 9.2 | - | - | 9.7, 13.7 |
| Patient death within 1 month of index, n (%) | <5 (0.4–1.6<sup>a</sup>) | <5 (1.4–5.5<sup>a</sup>) | 5 (3.8) | 58 (5.8) |
| 95% CI | - | - | 0.5, 7.1 | 4.3, 7.3 |

<sup>a</sup>Due to a total sample size of <5, this number was suppressed; a range of 1–4 events has been used to calculate the range given. CI, confidence interval.

### COVID-19-related hospitalisations during periods of BA.1, BA.2 and BA.5 predominance

During the BA.1 period, 281 patients were treated with sotrovimab, 68 with nirmatrelvir/ritonavir, 409 with molnupiravir and 2,742 patients received no treatment (Table 10). Similar to the overall analysis, a low proportion of patients treated with sotrovimab (n<5/281, 0.4–1.4%) experienced a COVID-19-related hospitalisation. Percentages for molnupiravir and untreated patients were 2.2% (95% CI 0.8%, 3.6%; n=9/409) and 3.2% (95% CI 2.5%, 3.9%; n=87/2,742), respectively. During the BA.2 period, there were 415 patients treated with sotrovimab, 269 treated with nirmatrelvir/ritonavir, 59 treated with molnupiravir and 1,302 who did not receive any treatment (Table 10). As for the BA.1 period, a low proportion of patients treated with sotrovimab (0.2–1.0%, n<5/415) experienced a COVID-19-related hospitalisation during the BA.2 period of prevalence. Molnupiravir-treated (1.7–6.8%, n<5/59) and untreated patients (2.1% [95% CI 1.3%, 2.9%], n=27/1,302) proportions were similarly low. It should be noted that as the total number of COVID-19-related hospitalisations in the nirmatrelvir/ritonavir cohort was fewer than 5, stratification in the BA.1 and BA.2 periods was not possible due to suppression rules.

**Table 10.** COVID-19-related hospitalisations during periods of BA.1 and BA.2 predominance

|  | **Sotrovimab**<br>**(n=696)** | **Nirmatrelvir/**<br>**ritonavir**<br>**(n=337)** | **Molnupiravir**<br>**(n=470)** | **Untreated**<br>**(n=4,044)** |
| --- | --- | --- | --- | --- |
| **Omicron BA.1 period** |  |  |  |  |
| N | 281 | 68 | 409 | 2,742 |
| COVID-19-related hospitalisation, n, % | <5 (0.4–1.4<sup>a</sup>) | - | 9 (2.2) | 87 (3.2) |
| 95% CI | - | - | 0.8, 3.6 | 2.5, 3.9 |
| **Omicron BA.2 period** |  |  |  |  |
| N | 415 | 269 | 59 | 1,302 |
| COVID-19-related hospitalisation, n (%) | <5 (0.2–1.0<sup>a</sup>) | - | <5 (1.7–6.8<sup>a</sup>) | 27 (2.1) |
| 95% CI | - | - | - | 1.3, 2.9 |

Due to the overall number of COVID-19-related hospitalisations being <5, stratification by BA.1 and BA.2 period could not be reported. <sup>a</sup>Due to a total sample size of <5, this number was suppressed; a range of 1–4 events has been used to calculate the range given. CI, confidence interval.

In a *post-hoc* exploratory analysis of BA.5 variant predominance during June and July 2022, Omicron BA.5 represented 70.6% of sequenced cases, with BA.2 and BA.4 representing 11.4% and 16.9% of sequenced cases, respectively.^26^ We found that 197 patients were treated with sotrovimab during this period, of whom fewer than five (0.5–2.0%) had a COVID-19-related hospitalisation (Table 11). Of 228 patients treated with nirmatrelvir/ritonavir, no patients (0.0%) experienced a COVID-19-related hospitalisation. In addition, fewer than five patients treated with molnupiravir experienced a COVID-19-related hospitalisation (7.7–30.8%; n<5/13). Lastly, of 657 patients who received no treatment, 1.8% (95% CI 0.8%, 2.8%; n=12) experienced a COVID-19-related hospitalisation.

**Table 11.** COVID-19-related hospitalisations during period of BA.5 predominance (*post-hoc* analysis)

|  | **Sotrovimab**<br>**(n=197)** | **Nirmatrelvir/**<br>**ritonavir (n=228)** | **Molnupiravir**<br>**(n=13)** | **Untreated**<br>**(n=657)** |
| --- | --- | --- | --- | --- |
| COVID-19-related hospitalisation, n (%) | <5 (0.5–2.0<sup>a</sup>) | 0 (0.0) | <5 (7.7–30.8<sup>a</sup>) | 12 (1.8) |
| 95% CI | - | - | - | 0.8, 2.8 |

<sup>a</sup>Due to a total sample size of <5, this number was suppressed; a range of 1–4 events has been used to calculate the range given.

## Discussion

The results of this study outline the characteristics and outcomes of patients who received early treatment for COVID-19 in the UK, or those who may have been eligible but did not receive treatment. Our results indicate that the characteristics of patients treated with the various early treatments are different, with evidence that sotrovimab is utilised amongst older and more at-risk patients.

Highest- and high-risk conditions were identified using SNOMED and ICD-10 codes from all available patient history. An unexpectedly large proportion of the treated cohorts had no evidence of the highest-risk conditions which make patients eligible for early COVID-19 treatment (sotrovimab: 39.2%, nirmatrelvir/ritonavir: 41.8%, molnupiravir: 45.7%). When these patient records were further evaluated, we found that a large proportion had an active outpatient appointment for oncology, renal, haematology, gastroenterology, or rheumatology services (sotrovimab: 54.2%, nirmatrelvir/ritonavir: 35.5%, molnupiravir: 47.9%). The majority of these patients also had the presence of a SNOMED code primarily used to identify patients who required shielding (a generic high-risk code shown via the Discovery dataset analysis to be predominantly used in the period during which shielding was introduced), thus indicating they were at high risk of developing COVID-19-related complications at some time during the pandemic (sotrovimab: 70.0%, nirmatrelvir/ritonavir: 62.4%, molnupiravir: 72.1%). This indicates that while these patients did not meet the case definition of highest-risk used in this study *per se*, they likely did have one or more underlying conditions that increased their risk of developing COVID-19-related complications and, therefore, would have been eligible for early treatments at an earlier time (aligned to national guidelines^8^).

Amongst the treated cohorts, the proportion of COVID-19-related hospitalisations was particularly low for sotrovimab-treated patients (0.7%) and nirmatrelvir/ritonavir-treated patients (0.3–1.2%). In total, 2.1% of molnupiravir-treated patients experienced a COVID-19-related hospitalisation, although it should be noted that the vast majority of patients receiving molnupiravir treatment (n=409/470) received treatment during the earlier Omicron BA.1 period. In addition, 2.8% of untreated patients experienced a COVID-19-related hospitalisation across the entire study period. Higher percentages of all-cause hospitalisations were reported across all cohorts compared with COVID-19-related hospitalisations. A similarly low proportion of patients with advanced renal disease treated with sotrovimab experienced severe outcomes due to COVID-19 (COVID-19-related hospitalisation and death within 1 month of index), although there was some evidence of worse outcomes for untreated patients. COVID-19-attributable hospitalisations appeared numerically consistent across age groups (18–64 and ≥65 years), as well as across the Omicron BA.1, BA.2 and BA.5 periods for the treated cohorts. However, formal statistical comparison adjusting for underlying patient characteristics is needed to confirm these results. There was some evidence of a reduction in the proportion of untreated patients with a COVID-19-related hospitalisation across the three time periods (3.2% during Omicron BA.1, 2.1% during BA.2 and 1.8% during Omicron BA.5). Potential reasons for this could include a reduction in the severity of the circulating variants^28^ or reduced immune naivety/susceptibility due to the UK’s vaccination programme or prior infection. There is also the possibility that with increased uptake of treatments, those patients most at risk were receiving treatment and therefore included in the treated cohorts, meaning that those remaining untreated were at lower risk of developing severe COVID-19. For example, on 24 December 2021, NHS guidance was updated to advise that only eligible patients whose symptoms had not shown signs of resolving should be offered treatment.^29^ Due to the differences in the underlying populations across each of the cohorts, no inferences can be made regarding the relative treatment benefits as part of this descriptive study. This will be the subject of a comparative effectiveness analysis adjusting for patient characteristics and potential other cofounders, which will be published subsequently.

There were also differences in participant characteristics between the treated cohorts included in this study and the populations of patients recruited for pivotal trials for each of the treatments. For example, the **CO**VID-19 **M**onoclonal Antibody **E**fficacy **T**rial-**I**ntent to **C**are **E**arly (COMET-ICE) randomised clinical trial, which compared sotrovimab with placebo,^13^ required patients have ≥1 risk factor for progression to severe COVID-19, including obesity, diabetes, congestive heart failure, chronic kidney disease and age ≥55 years. Of note, our sotrovimab-treated cohort had substantially more Asian/Asian British patients (23.9% vs 4.5% in COMET-ICE), while the COMET-ICE population included more White patients. In addition, our cohort included more patients with chronic kidney disease (20.4% vs 0.9% in COMET-ICE) and the COMET-ICE population included more patients with obesity (62.5% vs 2.3% in our sotrovimab-treated cohort), although it should be noted that obesity is often underreported in administrative data.

Although this study has not been designed to compare cohorts, there is evidence that a higher proportion of Black/Black British patients were untreated compared with those who received one of the early treatments investigated. These data highlight the ongoing need to support access for all patients eligible for such treatments, particularly when considering the association between social inequalities and COVID-19 morbidity, which is aggravated even further in the context of underlying comorbidities.^30^

The results of this study compliment other recent real-word studies on early treatments for COVID-19 in the UK.^31-33^ For example, the findings of a recent publication on the comparative effectiveness of sotrovimab versus molnupiravir in England, conducted using the OpenSAFELY platform, showed that sotrovimab treatment was associated with a significant reduction in risk of severe COVID-19 outcomes in comparison to treatment with molnupiravir during BA.1 (primary analysis) and BA.2 (exploratory analysis) predominance.^31^ Moreover, sotrovimab was found to offer a similar level of protection against disease progression in patients with COVID-19 infected with the Omicron BA.1 and BA.2 subvariants.^32^ Similarly, in an observational study in the US, treatment with sotrovimab was associated with a reduced risk of all-cause hospitalisation and mortality during the predominance of early Omicron variants.^33^

This study has several limitations that should be considered. Firstly, the study presents descriptive analyses with no adjustments for differences in patient characteristics between cohorts, meaning that any potential associations may be subject to bias and confounding. This will be addressed as part of a future comparative effectiveness analysis. Secondly, the severity of symptoms at baseline has not been captured as part of this study. This may affect treatment decisions as patients who have reducing symptoms are unlikely to be treated. In addition, the Discover dataset includes patients from a geographically restricted region (NWL), and therefore sub-national variation in treatment practices or patient demographics cannot be described. It should also be noted that, as for any real-world database study, ascertainment of positive COVID-19 cases and patient characteristics, including high-risk comorbidities, is reliant upon healthcare encounters and reliable administrative data recording, and missing data cannot therefore be ruled out. Also, as there were no sequencing data available for patients included in the study, we used dominance period for Omicron sub-variant as a surrogate.^26^ Finally, as with many analyses of electronic health records, the absence of a recorded diagnosis code is assumed to be synonymous with the absence of the disease itself.

## Conclusions

There is evidence of differences in the characteristics and risk profiles of patients with COVID-19 who are treated with sotrovimab, nirmatrelvir/ritonavir and molnupiravir, as well as eligible untreated patients. Sotrovimab was observed to be utilised frequently amongst patients with high-risk comorbidities, such as advanced renal disease, which may place patients at increased risk of severe COVID-19. The proportion of patients with COVID-19-related hospitalisations was particularly low for sotrovimab- and nirmatrelvir/ritonavir-treated patients, with findings for sotrovimab being maintained amongst patients with advanced renal disease, as well as patients aged both 18–64 and ≥65 years. Across the Omicron BA.1, BA.2 and BA.5 periods (through July 2022), COVID-19 hospitalisations for all treated cohorts were consistently low.

## Data Availability

The Discover-NOW data that support the findings of this study are available from Imperial College Health Partners via approval from the Discover Data Access Group (DRAG) under certain restrictions.

## Acknowledgements

Editorial support (in the form of writing assistance, including preparation of the draft manuscript under the direction and guidance of the authors, collating and incorporating authors’ comments for each draft, assembling tables and figures, grammatical editing and referencing) was provided by Mirela Panea and Kathryn Wardle of Aura, a division of Spirit Medical Communications Group Limited (Manchester, UK), and was funded by GSK.

## Declarations

### Ethics statement

This study was approved by the Discover Data Access Group (DRAG) which has derogated powers to approve studies under its overarching approval as a Health Research Database. This study complies with all applicable laws regarding subject privacy. Data were aggregated and counts less than five were suppressed in line with IG suppression rules. No direct subject contact or primary collection of individual human subject data occurred. Study results were in tabular form and aggregate analyses that omit subject identification, therefore informed consent, ethics committee or IRB approval were not required. Any publications and reports do not include subject identifiers.

### Author contributions

All authors made a significant contribution to the work reported, whether that was in the conception, study design, execution, acquisition of data, analysis and interpretation, or in all these areas; took part in drafting, revising or critically reviewing the manuscript; gave final approval of the version to be published; have agreed on the journal to which the article has been submitted; and agree to be accountable for all aspects of the work.

