## Supplemental Material for "Characteristics and outcomes of patients with COVID-19 at high-risk of disease progression receiving sotrovimab, oral antivirals or no treatment in England"

### **Table S1.** Source datasets for key study data items in the DISCOVER-NOW data warehouse

| **Data items** | **Source** |
| --- | --- |
| COVID-19 diagnosis data | North-West London pathology dataset |
| Anti-COVID-19 drug prescriptions (and other prescriptions of interest; agents to identify highest-risk patients) | Sotrovimab only, high-cost drugs dataset (Bluteq)  Chelsea & Westminster hospitals pharmacy datasets (available via linkage to pharmacy data)  Primary care datasets |
| Patient demographic & clinical characteristics | Primary care datasets |
| HCRU | Secondary care data |

COVID-19, coronavirus disease 2019; HCRU, healthcare resource utilisation.
